# Insights from a Decade of Hereditary Cancer Genetic Testing from India: A Retrospective Study

**DOI:** 10.64898/2026.07.29.26358035

**Authors:** Ramesh Menon, Lakshmi Mahadevan, Aviral Kumar, Akshi Bassi, Lavina Udhwani, Ankit Verma, Anurag Gupta, Lavanya Balakrishnan, Manju Lakshmi, Priyanka Shrivastav, Anisha Tomy, Aarya Senan, Meghana Raman, Thenral S. Geetha, Greeshma Purushothama, Hamsini Koduru, Charu Bahl, Anand Pathak, Bharath Rangarajan, Ram Abhinav, Firoz Rajan, Somnath Roy, Anik Ghosh, Joydeep Ghosh, Jaydip Bhaumik, Arnab Bhattacharjee, Sanjit Agrawal, Priya Tiwari, Akash Tiwari, Nahush Tahiliani, Shona Nag, Arun Warrier, Aju Mathew, Alec Reginald Errol Correa, Ajay Bapna, Harsh Sheth, Sachin Hingmire, Neelesh Reddy, Deepak Shukla, Paul Augustine, Bhuvan Chugh, Sankar Srinivasan, Chetna Bakshi, Abdul Shahid, Amit Rauthan, Yogesh Mistry, Prasanth Parameswaran, Senthil J Rajappa, Sanju Cyriac, Asima Mukhopadhyay, Harit Chaturvedi, Aparna Dhar, Pramod Kumar Julka, Atul Batra, Raja Pramanik, Gomathi Shankar, Bhargavi Ilangovan, Rajiv Sarin, Sakthivel Murugan, Ramprasad L Vedam, Ravi Gupta

## Abstract

**Background:** Hereditary cancers account for approximately 5% to 10% of malignancies, with inherited susceptibility more frequently identified in individuals with early-onset disease or a significant family history. However, large-scale study of germline cancer susceptibility across multiple tumor types remains limited from India and the broader South Asian population.

**Methods:** We retrospectively analyzed 24,320 individuals who underwent germline hereditary cancer testing at MedGenome Labs Ltd., Bangalore, India, between 2016 and 2025. Testing was performed primarily using clinically validated multi-gene targeted next-generation sequencing (NGS) panels (N = 22,773), and for a subset of these cases were analyzed using multiplex ligation-dependent probe amplification (MLPA) (N = 4,524). Additionally, MLPA confirmatory test alone was done for 1,547 samples. The major cancer subgroups analyzed were breast (N = 10,292), ovarian (N = 3,935), colorectal (N = 1,267), prostate (N = 762), and endometrial (N = 539) cancers, with 2,764 asymptomatic individuals with a positive family history.

**Results:** The overall diagnostic yield was 23.9%, highest in colorectal (42.1%) followed by ovarian (31.6%), endometrial (22.3%), breast (20.3%), and prostate (8.7%) cancers. Family history of the corresponding cancer was associated with higher diagnostic yield across most major cancer types, whereas prostate cancer showed a higher yield among individuals reporting a family history of other cancers. Diagnostic yield generally decreased with increasing age in breast, colorectal, ovarian, and endometrial cancers, but increased with age in prostate cancer. *BRCA1* and *BRCA2* predominated in breast and ovarian cancers, mismatch repair genes in colorectal and endometrial cancers, and *BRCA2* in prostate cancer. The recurrent *BRCA1* c.68_69delAG (p.Glu23ValfsTer17) variant was identified in 351 individuals. The identified *BRCA1* gene mutations correlated with functional consequence data from published mutagenesis study. Among breast cancer cases, the ER−/HER2− subtype showed the highest diagnostic yield (25.5%). Re-analysis of VUS using *autoACMG* classifier followed by manual review confirmed 277 of 443 samples (62.5%) as pathogenic/likely pathogenic (P/LP). Of the remaining VUS, 3.2% were classified as VUS-High using *VUS-Grader* tool. Additionally, manual review of 74 CNV-positive samples classified as VUS resulted in the reclassification of 41 (55.4%) as likely pathogenic, increasing the diagnostic yield by 1.4 percentage points. Among 4,524 individuals who underwent combined NGS and MLPA testing, CNVs were identified in 251 (5.5%), including 59 cases detected by MLPA but not by NGS.

**Conclusions:** This retrospective study represents the largest characterization of hereditary cancer susceptibility across major cancer types in South Asia, encompassing 24,320 samples. The findings support comprehensive multi-gene testing and population-adapted testing strategies to improve hereditary cancer risk assessment and precision oncology.

## Introduction

Hereditary cancers arise from germline pathogenic variants in cancer predisposition genes^1^. Identification of these pathogenic variants enables risk stratification, cascade testing, targeted surveillance, and risk-reducing strategies^1,2^. Advances in next-generation sequencing have transformed hereditary cancer testing from single-gene assays to multigene panels, enabling simultaneous assessment of genes involved in homologous recombination repair, mismatch repair, and other cancer predisposition pathways^3^. Germline testing recommendations have increasingly expanded across cancer types, with growing support for broader testing beyond traditional age- and family-history–based criteria^4^.

The expansion of multigene testing has increased the number and diversity of variants detected, creating a greater need for standardized variant interpretation. Variant classification follows criteria established by the American College of Medical Genetics and Genomics and the Association for Molecular Pathology^5^. Interpretation also relies on curated databases such as ClinVar^6^ and population allele frequency resources such as the 1000 Genomes^7^, Genome Aggregation Database^8^, TopMed^9^, and GenomeAsia^10^. Despite expanding testing capabilities, the increased detection of variants, particularly variants of uncertain significance (VUS), continues to pose interpretive challenges^11^.

South Asian populations remain underrepresented in global genomic reference datasets, contributing to greater diagnostic uncertainty and potentially higher VUS rates^12–15^. India faces a growing cancer burden, with breast cancer as the most common malignancy among women and rising incidence of colorectal and prostate cancers^16,17^. Studies of germline testing in breast and ovarian cancers in India have generally been limited to relatively small, disease-specific cohorts^18–22^. Large-scale, pan-cancer studies integrating multigene germline testing with copy number variant (CNV) detection remain limited in India and the broader South Asian population.

In this study, we conducted a retrospective analysis of 22,773 individuals who underwent germline hereditary cancer testing at MedGenome Labs Ltd., one of the largest genetic testing laboratories in India. The cohort included individuals with breast, ovarian, colorectal, prostate and endometrial cancers, as well as other cancer types. Asymptomatic individuals with a positive family history were analyzed as a separate group. Germline testing was performed using multigene panels ranging from BRCA1/2-focused panels to comprehensive panels of up to 158 genes, with a subset of individuals also evaluated using multiplex ligation-dependent probe amplification (MLPA) for CNV detection. The objectives of this study were to determine overall and cancer-specific diagnostic yields; characterize age, sex, and family-history distributions; examine clinical factors associated with germline findings; characterize gene-specific variant spectra; evaluate breast cancer hormonal receptor subtypes and assess the contribution of MLPA-based CNV detection. By analyzing one of the largest hereditary cancer datasets reported from South Asia, this study provides real-world evidence on hereditary cancer susceptibility in an underrepresented population.

## Materials and Methods

### Study design and patient cohort

This retrospective study analyzed germline hereditary cancer testing data from MedGenome Laboratories, Bangalore, India. The comprehensive dataset comprised 22,773 unique individuals who underwent germline next-generation sequencing (NGS)-based hereditary cancer testing over a 10-year period, from 2016 to 2025. Clinical and demographic variables were obtained from the Test Requisition Form (TRF) which included age at testing, sex, cancer type, family history, metastatic status, NGS panel type, and variant classification. Family history was defined as the presence of the same or related cancers in first- or second-degree relatives. Based on the information provided by the referring clinician, individuals were classified according to cancer indication into the following categories: breast (n = 10,292), ovarian (n = 3,935), colorectal (n = 1,267), prostate (n = 762), endometrial (n = 539), other cancer types (n = 3,214), and asymptomatic individuals with a reported family history of cancer (n = 2,764).

### Targeted germline multigene panel sequencing

Genomic DNA extracted from peripheral blood samples was subjected to germline analysis using four clinically validated gene panels: (i) a comprehensive Hereditary Cancer Panel (HCP) comprising 158 genes; (ii) a germline *BRCA1*/*BRCA2* (gBRCA) panel including 2 genes; (iii) a Hereditary Breast and Ovarian Cancer (HBOC) panel comprising 39 genes; and (iv) a germline Homologous Recombination Repair (gHRR) panel comprising 15 genes.

A total of 50ng of high-quality genomic DNA was used for library preparation employing clinically validated DNA library preparation kits in accordance with manufacturer protocols. Target enrichment was performed using panel-specific custom hybrid-capture probes. The enriched libraries were subjected to paired-end sequencing (read length of 150 bp) on Illumina platforms used during the study period, including the HiSeq 2500, HiSeq 4000, HiSeq X Ten, and NovaSeq X, all based on Illumina sequencing-by-synthesis technology.

### Copy number variation detection by Multiplex ligation-dependent probe amplification (MLPA)

CNV analysis was performed using MLPA in two groups of individuals: 4,524 individuals who underwent combined next-generation sequencing (NGS) and MLPA testing as part of the primary study cohort, and an additional 1,547 individuals who underwent MLPA testing alone. MLPA was used to detect clinically relevant exon-level deletions and duplications. The SALSA digitalMLPA Probemix NXtec D002 Hereditary Cancer Panel 2 (MRC Holland, Amsterdam, the Netherlands) was used for MLPA analysis.

### Variant interpretation

Sequencing reads were aligned to the human reference genome builds GRCh37 (hg19) or GRCh38 (hg38), depending on the bioinformatics pipeline version in use at the time of clinical report generation. Variant annotation was performed using the MedGenome in-house VariMAT tool, which uses Variant Effect Predictor (VeP) program^23^. Variants were annotated against ClinVar^6^, OMIM^24^, HGMD^25^, and filtered based on population allele frequencies from the 1000 Genomes Project^7^, ExAC^26^, gnomAD^8^, dbSNP^27^, the integrative Japanese genome variation database^28^, GenomeAsia^10^, and an internal database. Functional impact was assessed using PolyPhen-2^29^, SIFT^30^, MutationTaster2^31^, and LRT^32^. Variants were prioritized according to inheritance pattern and phenotype and classified following ACMG guidelines. The versions of the tools and databases are based on their updates from 2016 to 2025.

### Sample level variant classification and diagnostic yield

For sample-level variant classification, an individual could harbor variants from multiple classification categories (Pathogenic (P), likely pathogenic (LP), and/or variants of uncertain significance (VUS). For analyses requiring a single classification per sample, including calculation of diagnostic yield, priority was assigned to P, followed by LP and VUS. Thus, an individual harboring both LP and VUS variants was classified as LP, while an individual harboring a P variant was classified as P. Diagnostic yield was defined as the proportion of samples classified as P/LP among all samples undergoing germline testing. Accordingly, reported numbers represent samples rather than unique variants.

### Reclassification of samples with VUS variants

Variants of uncertain significance (VUS) identified as SNVs and indels were re-evaluated using an in-house ACMG-based classification tool (*autoACMG),* which automated 17 of the 28 ACMG rules to accurately classify and interpret variants in accordance with the guidelines. Variants classified as P/LP were subsequently reviewed manually to assess their clinical relevance based on patient phenotypes. As *autoACMG* does not support CNV classification, CNV-positive samples classified as VUS were evaluated separately through manual review for potential reclassification. When multiple reclassification outcomes were present within the same sample, the highest classification was used for sample-level analysis, with priority assigned to P, followed by LP and VUS. Accordingly, reported numbers represent samples rather than unique variants. Variants that remained classified as VUS after reclassification were further evaluated using an automated VUS-Rounds method^33^. Positive scores were assigned for evidence supporting pathogenicity, including prior P/LP reports, predicted deleterious effects, and functional evidence, whereas negative scores were assigned for evidence supporting benignity, including excessive population allele frequency, limited gene–disease validity, prior benign reports, or predicted tolerance based on a consensus of *in silico* predictors. The aggregate score (HotVUS score) was used to subclassify VUS as VUS-High (>2 to ≤7), VUS-Mid (>0 to ≤2), or VUS-Low (<0)^34^.

### Statistical analysis

A multi-variate logistic regression analysis was performed to evaluate factors associated with P/LP variant detection among individuals with a personal history of breast, ovarian, colorectal, endometrial, or prostate cancer. For this analysis, breast cancer was used as a reference following the approach of Papadopoulou et al^35^. Diagnostic yield was defined as detection of a P/LP germline variant according to ACMG/AMP classification; VUS and benign findings were considered non-diagnostic. Family history was categorized as none, concordant (history of the same cancer type as the patient), or discordant (history of a different cancer type only). The model included cancer type (reference, breast), family history (reference, none), and age as predictors. Age was modeled per 10-year increase and centered at 50 years. A second model included a cancer-type-by-age interaction to assess whether the association between age and diagnostic yield differed by cancer type. Results are reported as adjusted odds ratios (ORs) with 95% CIs. Analyses were performed using Python (version 3.12) and the statsmodels library (version 0.14.5).

## Results

### Cohort characteristics

A total of 22,773 unique individuals who underwent germline hereditary cancer testing were included in the study. The cohort showed broad geographic representation across India, with samples contributed by major cities including New Delhi, Kolkata, Mumbai, Bangalore, Chennai, Hyderabad, Ahmedabad, Puducherry, Thiruvananthapuram, Pune, Kochi, and Jaipur (**Fig. 1A**). Breast cancer was the predominant indication for testing (45.2%), followed by ovarian cancer (17.3%), colorectal cancer (5.6%), prostate cancer (3.4%), and endometrial cancer (2.4%). Other cancer types accounted for 14.1% of the cohort, while 12.1% comprised asymptomatic individuals with a reported family history of cancer (**Fig. 1B**). The cohort was predominantly female (83.1%), and approximately half of the individuals (51.5%) were aged ≤50 years (**Table 1** and **Fig. 1C–D**).

**Figure 1.**
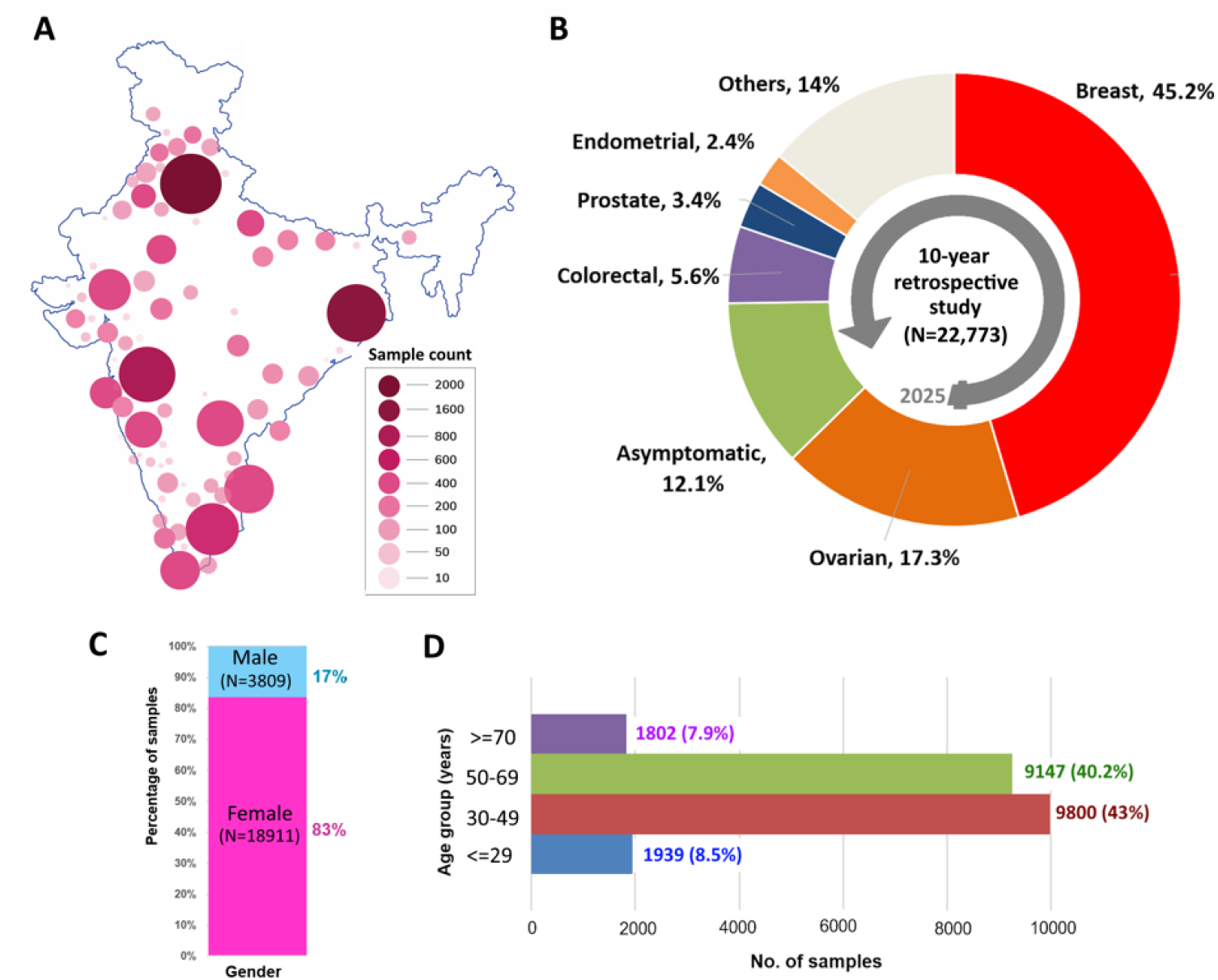
Demographic and clinical characteristics of the study cohort. **(A)** Geographic distribution of study participants across India. Circle size and colour intensity indicate the number of samples contributed from each location, with larger circles and darker shades representing higher sample counts. **(B)** Distribution of participants by clinical indication, including breast, ovarian, colorectal, prostate, and endometrial cancers, asymptomatic individuals, and other indications. **(C)** Sex distribution of the cohort, comprising 83.1% females (n = 18,911) and 16.7% males (n = 3,809); sex information was unavailable for 53 individuals (0.2%). **(D)** Distribution of participants across age groups (≤29, 30–49, 50–69, and ≥70 years). The x-axis represents the number of samples, and the y-axis represents age groups.

**Table 1.** Demographic characteristics of the study cohort across clinical categories.

| **Clinical category** | **Sample count (%)** | **Age group** |  |  |  |  | **Gender** |  |  |
|---|---|---|---|---|---|---|---|---|---|
|  |  | **≤29** | **30-49** | **50-69** | **≥70** | **Not known** | **Male (%)** | **Female (%)** | **Not known** |
| Breast Cancer | 10292 (45.2%) | 530 (5.1%) | 5445 (52.9%) | 3747 (36.4%) | 533 (5.2%) | 37 (0.4%) | 174 (1.7%) | 10100 (98.1%) | 18 (0.2%) |
| Ovarian Cancer | 3935 (17.2%) | 221 (5.6%) | 1276 (32.4%) | 2110 (53.6%) | 301 (7.6%) | 27 (0.7%) | 0 | 3935 (100%) | 0 |
| Colorectal Cancer | 1267 (5.6%) | 123 (9.7%) | 467 (36.9%) | 537 (42.4%) | 138 (10.9%) | 2 (0.2%) | 776 (61.2%) | 487 (38.4%) | 4 (0.3%) |
| Prostate Cancer | 762 (3.4%) | 1 (0.1%) | 16 (2.1%) | 410 (53.8%) | 334 (43.8%) | 1 (0.1%) | 762 (100%) | 0 | 0 |
| Endometrial Cancer | 539 (2.4%) | 10 (1.9%) | 119 (22.1%) | 350 (64.9%) | 60 (11.1%) | 0 | 0 | 539 (100%) | 0 |
| Asymptomatic individuals | 2764 (12.1%) | 477 (17.3%) | 1526 (55.2%) | 683 (24.7%) | 69 (2.5%) | 9 (0.3%) | 677 (24.5%) | 2069 (74.9%) | 18 (0.7%) |
| Others | 3214 (14%) | 577 (18.0%) | 951 (29.6%) | 1310 (40.8%) | 367 (11.4%) | 9 (0.3%) | 1405 (43.7%) | 1800 (56.0%) | 9 (0.3%) |

Age distribution varied across cancer groups. Breast cancer and the asymptomatic group had a greater representation of younger individuals, with 52.9% of individuals with breast cancer and 55.2% of asymptomatic individuals aged 30–49 years. In the asymptomatic group, 72.5% of individuals were aged <50 years, compared with 51.5% of the overall cohort (**Supp. Fig. 1A** and **Table 1**). Ovarian, colorectal, and endometrial cancers were most frequently observed among individuals aged 30–69 years, whereas prostate cancer was predominantly observed in individuals aged ≥70 years (43.8%) (**Table 1** and **Supp. Fig. 1A**). Females accounted for 98.1% of breast cancer cases, while males comprised 61.2% of colorectal cancer cases (**Table 1** and **Supp. Fig. 1B**).

### Variant distribution across cancer types

Four next-generation sequencing (NGS) panels were used for germline genetic testing: HCP, gBRCA, gHRR, and HBOC. The HCP panel accounted for the majority of tests (77.8%), followed by gBRCA (20.4%), gHRR (1.1%), and HBOC (0.6%). Across all panels, SNVs were the most frequently detected variant type, followed by indels and CNVs. Pathogenic (P) variants constituted the largest proportion of classified findings, while VUS findings were most frequent with the HCP panel (26.3%). The highest proportion of cases with no variants detected was observed with the gBRCA panel (73.5%).

The distribution of variant types varied across the five major cancer types. SNVs were predominant across all cancer groups, with the highest proportion observed in colorectal cancer (46.3%). Indels were the second most frequent variant type, while CNVs accounted for a smaller proportion, ranging from 0.5% in prostate cancer to 4.5% in colorectal cancer.

The distribution of variant classifications also differed across cancer types (**Fig. 2A**). Pathogenic (P) variants accounted for the highest proportion of findings in colorectal cancer (37%), followed by ovarian (29%), endometrial (19%), breast (18%), and prostate cancer (7%). VUS accounted for 17%–26% of findings across cancer types, while cases with no variants detected ranged from 32% in colorectal cancer to 69% in prostate cancer.

**Figure 2.**
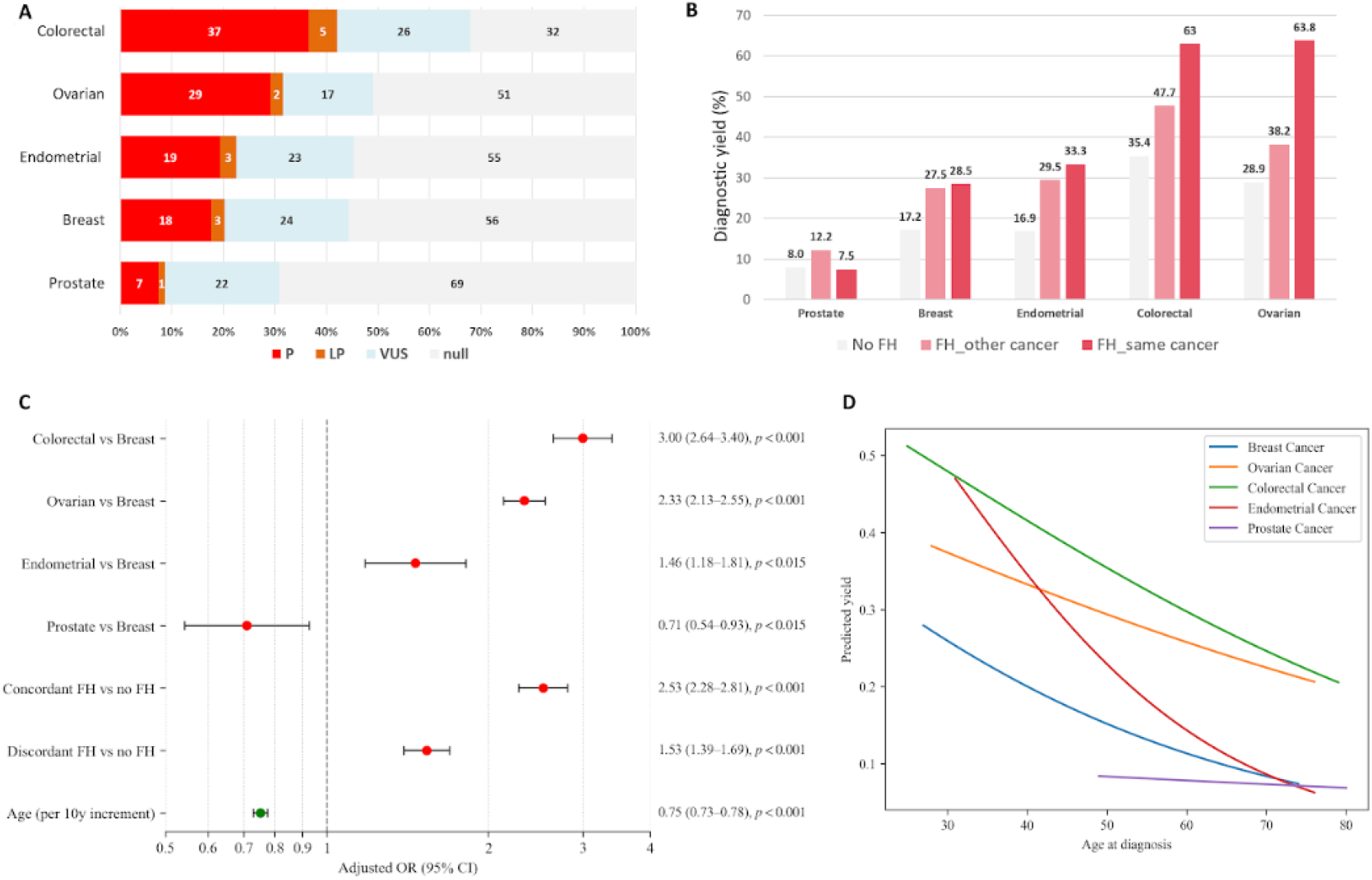
Variant distribution across cancer types, diagnostic yield and family history. **(A)** Distribution of variant classifications across colorectal, ovarian, endometrial, breast, and prostate cancers, categorized as pathogenic (P), likely pathogenic (LP), variants of uncertain significance (VUS), and cases with no variants detected (Null). The x-axis represents the percentage of individuals, and the y-axis represents cancer types. **(B)** Diagnostic yield (%) stratified by family history status (no family history, family history of another cancer type, and family history of the same cancer type). The x-axis represents cancer types, and the y-axis represents diagnostic yield (%). **(C)** Adjusted odds ratios (ORs) with 95% confidence intervals (CIs) and p-values from the multi-variate logistic regression model, with breast cancer as the reference category. The outcome was detection of a P/LP variant, and age was modeled as a continuous co-variate per 10-year increment. **(D)** Predicted diagnostic yield by age at diagnosis and cancer type, based on a logistic regression model incorporating an interaction between age and cancer type.

### Diagnostic yield and associated clinical factors

The overall diagnostic yield across the study cohort was 23.9%. Cancer-specific diagnostic yields varied across indications, with the highest yield observed in colorectal cancer (42.1%), followed by ovarian cancer (31.6%), endometrial (22.3%), breast (20.3%), and prostate cancer (8.7%). Among asymptomatic individuals with a reported family history of cancer, the diagnostic yield was 19.1%. The diagnostic yields observed for breast and/or ovarian cancers were comparable to those reported in previous Indian studies, which ranged between 18.6% to 30.1%^18–21,37,38^.

The age-wise distribution of diagnostic yield across the five major cancer types is presented in **Supp. Fig. 2**. Diagnostic yield generally decreased with increasing age across colorectal, ovarian, endometrial, and breast cancers. In contrast, prostate cancer showed an increasing trend in diagnostic yield with age, with the highest yield observed among individuals aged ≥70 years (10.4%).

For breast cancer, diagnostic yield was also evaluated among individuals diagnosed at <50 years of age, consistent with an age-based criterion for hereditary cancer testing in National Comprehensive Cancer Network (NCCN) guidelines. The diagnostic yield in this subgroup was 24.3%, slightly higher than that observed in the overall breast cancer cohort.

Family history patterns varied across cancer types. A family history of the same cancer type was most frequently reported among individuals with colorectal (11%) and breast cancer (10%), followed by prostate (5%), endometrial (3%), and ovarian cancer (2%). Conversely, a substantial proportion of individuals with prostate (78%) and ovarian cancer (77%) reported no family history of cancer. Endometrial cancer had the highest proportion of individuals reporting a family history of other cancer types (39%) (**Supp. Fig. 3**). Among asymptomatic individuals with a reported family history of cancer, breast cancer was the most frequently reported cancer in the family history (28.4%), followed by ovarian (7.4%) and colorectal cancer (4.6%).

**Figure 3.**
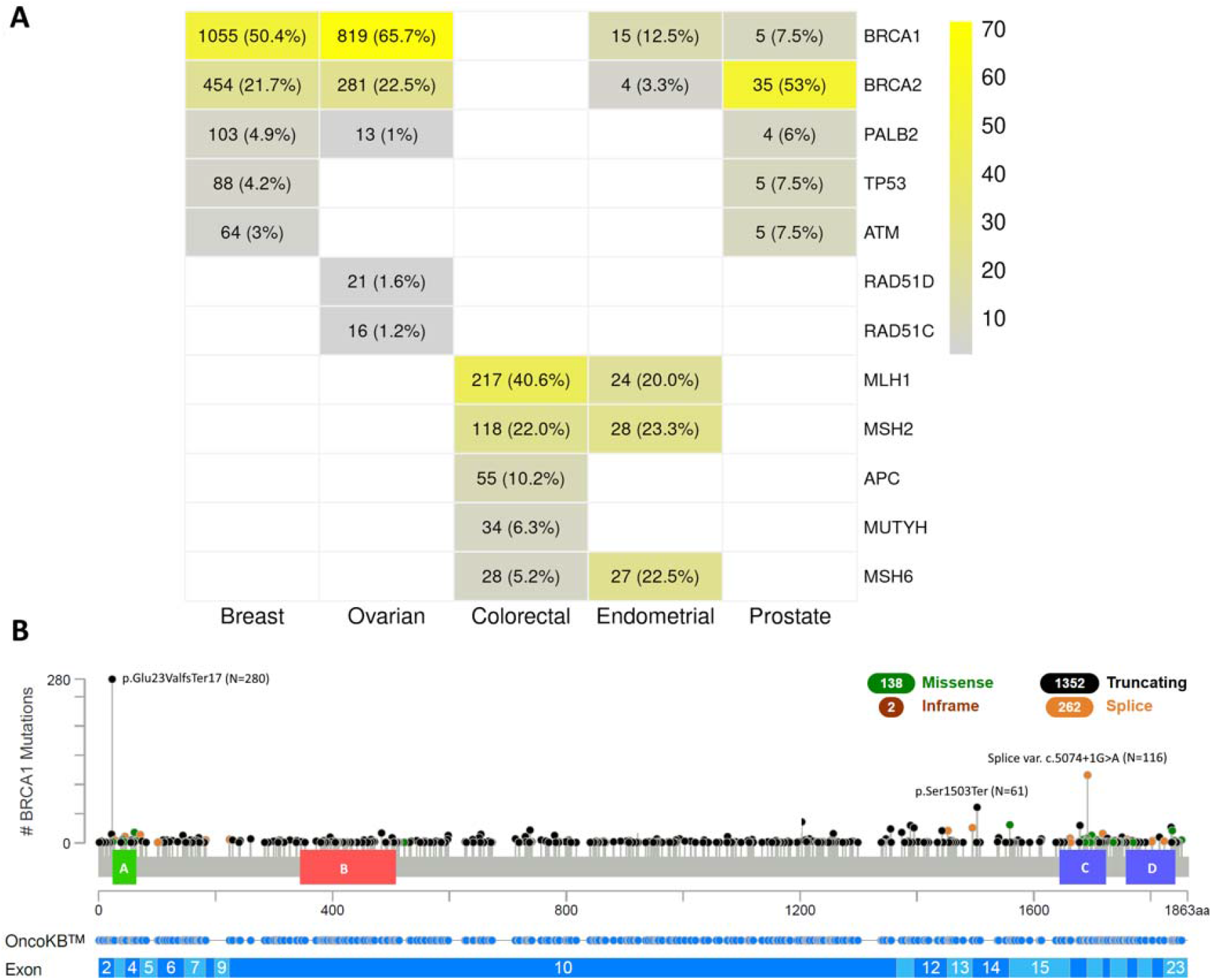
Cancer-specific susceptibility gene profiles and recurrent BRCA1 pathogenic variants in breast and ovarian cancers. **(A)** Most frequently affected genes in breast, ovarian, colorectal, endometrial, and prostate cancers, showing the number and percentage of P/LP variants within each cancer type. **(B)** Lollipop plot showing the three most frequently detected recurrent *BRCA1* pathogenic variants in the combined breast and ovarian cancer cohort. Variant labels, variant counts and positions are displayed as generated by the lollipop plot tool. The annotated protein domains are: A, zf-C3HC4 zinc-finger (RING finger) domain (amino acids 24–64); B, BRCT-associated serine-rich domain (amino acids 345–508); C, BRCA1 C-terminal (BRCT) domain (amino acids 1644–1723); and D, BRCA1 C-terminal (BRCT) domain (amino acids 1758–1842).

Diagnostic yield was further stratified by family history status (**Fig. 2B**). Across cancer types, individuals with a family history of the same cancer generally had the highest diagnostic yields compared with those reporting a family history of other cancers or no family history. Among individuals with ovarian cancer, the diagnostic yield was 63.8% in those with a family history of ovarian cancer, compared with 38.2% in those with a family history of other cancers and 28.9% in those without a family history. Similar patterns were observed for colorectal, endometrial, and breast cancers. In contrast, among individuals with prostate cancer, the diagnostic yield was slightly higher in those with a family history of other cancers than in those with a family history of prostate cancer.

Diagnostic yield was also compared between metastatic and non-metastatic cases across the five major cancer types. Non-metastatic cases showed higher diagnostic yields than metastatic cases in breast (20.6% vs. 18.2%), colorectal (43.7% vs. 25.7%), endometrial (22.7% vs. 17.6%), and prostate cancers (9.1% vs. 7.9%). In contrast, metastatic ovarian cancer cases showed a higher diagnostic yield than non-metastatic cases (34.6% vs. 31.3%).

In the multi-variate logistic regression model, with breast cancer as the reference category, the odds of detecting a pathogenic or likely pathogenic (P/LP) variant were higher among individuals with colorectal cancer (OR 3.00, 95% CI 2.64–3.40), ovarian cancer (OR 2.33, 95% CI 2.13–2.55), and endometrial cancer (OR 1.46, 95% CI 1.18–1.81). In contrast, individuals with prostate cancer had lower odds of P/LP detection (OR 0.71, 95% CI 0.54– 0.93) (**Fig. 2C**).

Family history was also associated with P/LP detection. Compared with individuals without a reported family history of cancer, those with a family history of the same cancer type had more than twice the odds of P/LP detection (OR 2.53, 95% CI 2.28–2.81), while those with a family history of a different cancer type had a smaller increase in odds (OR 1.53, 95% CI 1.39–1.69). Age was independently associated with diagnostic yield, with the odds of P/LP detection decreasing by approximately 25% for every 10-year increase in age (OR 0.75, 95% CI 0.73–0.78) (**Fig. 2C**).

Although the observed diagnostic yield increased with age in prostate cancer, model-based predicted yields decreased with increasing age across all five cancer types in the logistic regression model incorporating an age-by-cancer-type interaction (**Fig. 2D**). The magnitude of this decline varied by cancer type. Endometrial cancer showed the steepest decline in predicted yield, whereas colorectal cancer maintained the highest predicted yield across most of the age range. Ovarian cancer showed a more gradual decline, while breast cancer demonstrated a progressive decrease in predicted yield with age. Prostate cancer had the lowest predicted yield across the age range and showed comparatively little change with increasing age.

### Top susceptibility genes and recurrent *BRCA1* variants

The most frequently implicated susceptibility genes across the five major cancer types are summarized in **Fig. 3A**. Analysis of P/LP variants revealed distinct gene-specific profiles across cancer types. In breast cancer, *BRCA1* was the most frequently affected gene, accounting for 50.4% of findings, followed by *BRCA2* (21.7%), *PALB2* (4.9%), *TP53* (4.2%), and *ATM* (3.0%). A similar pattern was observed in ovarian cancer, where BRCA1 accounted for 65.7% of findings, followed by *BRCA2* (22.5%) and other homologous recombination repair genes, including *RAD51D* (1.6%), *RAD51C* (1.2%), and *PALB2* (1.2%).

In contrast, colorectal and endometrial cancers showed a predominance of mismatch repair genes. In colorectal cancer, *MLH1* was the most frequently affected gene (40.6%), followed by *MSH2* (22.0%), *APC* (10.2%), *MUTYH* (6.3%), and *MSH6* (5.2%). Similarly, endometrial cancer showed a high frequency of P/LP variants in *MSH2* (23.3%), *MSH6* (22.5%), and *MLH1* (20.0%), with additional contributions from *BRCA1* (12.5%) and *BRCA2* (3.3%). In prostate cancer, *BRCA2* was the most frequently affected gene (53.0%), followed by *TP53*, *BRCA1*, and *ATM* (7.5% each), and *PALB2* (6.0%).

Given the predominance of *BRCA1* in breast and ovarian cancers, recurrent *BRCA1* pathogenic variants were further examined in the combined breast and ovarian cancer cohort (n = 14,227). The most frequently detected variant was c.68_69del (p.Glu23ValfsTer17), an Ashkenazi Jewish founder variant, identified in 1.97% of the cohort. This was followed by the splice-site variant c.5074+1G>A (0.64%), c.4508C>A (p.Ser1503Ter) (0.26%), c.3607C>T (p.Arg1203Ter) (0.25%), and c.4165_4166delAG (p.Ser1389Ter) (0.21%). The three most frequently detected recurrent *BRCA1* variants are highlighted in the lollipop plot in **Fig. 3B**. All five variants were observed more frequently in breast cancer than in ovarian cancer cases.

### Functional evaluation of the *BRCA1* variants

To further evaluate the functional relevance of *BRCA1* variants detected in this study, we compared our variant classifications with the functional categories reported in the BRCA1 saturation mutagenesis dataset by Findlay et al.^36^ This study categorized variants into loss-of-function (LoF), intermediate, and functional (FUNC) classes. LoF variants severely disrupt BRCA1 protein activity, intermediate variants partially impair function, and FUNC (neutral/wild-type–like) variants show little or no measurable effect on protein function. Among the 3,893 *BRCA1* variant positions reported in the mutagenesis dataset, 152 variants identified in our cohort overlapped with these positions (3.90%). The majority of overlapping variants were classified as LoF (117 variants), followed by FUNC (27 variants) and intermediate variants (8 variants). Consistent with functional expectations, most pathogenic and likely pathogenic (P/LP) variants identified in our cohort corresponded to LoF variants reported in the mutagenesis study, including 81 pathogenic and 19 likely pathogenic variants within the LoF category. In contrast, variants categorized as functional (FUNC) in the mutagenesis dataset were predominantly classified as variants of uncertain significance (VUS) in our analysis, accounting for 25 of the 27 FUNC variants.

### Distribution and diagnostic yield of breast cancer hormone receptor-based sub-types

Among the 5,234 breast cancer cases with available hormone receptor subtype information, ER−/HER2− was the predominant subtype (59.2%), followed by ER+/HER2− (29.5%), ER+/HER2+ (6.6%), and ER−/HER2+ (4.7%). Diagnostic yield varied across subtypes, with the highest yield observed in the ER−/HER2− subgroup (25.7%) and the lowest in the ER−/HER2+ subgroup (10.2%). The observed-to-expected (O/E) ratio was also highest in the ER−/HER2− subgroup (1.3), compared with 0.8 for both ER+/HER2− and ER+/HER2+ and 0.5 for ER−/HER2+ (**Supp. Fig. 4**).

**Figure 4.**
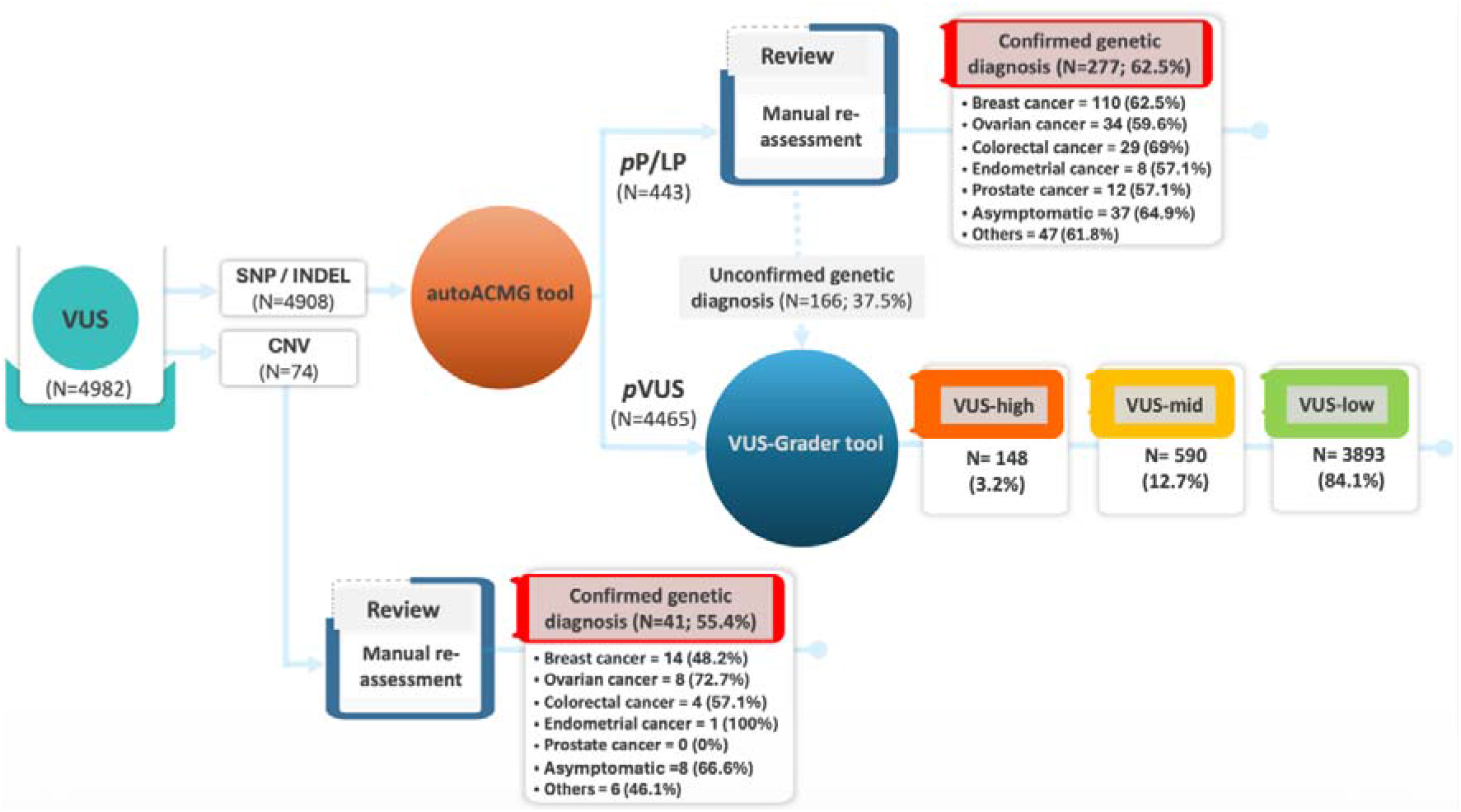
Workflow for reclassification of variants of uncertain significance (VUS). VUS-positive samples carrying SNVs and indels were reanalysed using the *autoACMG* tool, followed by manual review of samples classified as P/LP to assess their clinical relevance. Samples that remained classified as VUS were further stratified into VUS-High, VUS-Mid, and VUS-Low categories using the *VUS Grader* tool. CNV-positive samples classified as VUS were evaluated separately through manual review for potential reclassification.pP/LP, predicted pathogenic/likely pathogenic; pVUS, predicted variant of uncertain significance.

### Re-assessment of samples with uncertain genetic diagnosis

To further refine the variant interpretation, samples with VUS variants were reanalysed using the reclassification strategy illustrated in **Fig. 4**. A total of 4,908 SNV/indel-positive samples were reassessed using ACMG classifier tool (*autoACMG*), resulting in the reclassification of 443 samples as pathogenic or likely pathogenic (P/LP). Following manual review, 277 (62.5%) samples were confirmed for P/LP variants based on their clinical phenotypes, while the remaining 166 (37.5%) continued to be classified as VUS (**Fig. 4**). The proportion of samples reclassified as P/LP ranged from 57.1% in prostate and endometrial cancers to 69.0% in colorectal cancer. Among the asymptomatic individuals, 37 of 57 samples (64.9%) were confirmed as P/LP, while among individuals with other cancer types, 47 of 76 samples (61.8%) were confirmed as P/LP.

The 4,631 samples that remained classified as VUS following *autoACMG* reanalysis and manual review were further assessed using the *VUS-Grader* tool and categorized as VUS-High (148; 3.2%), VUS-Mid (590; 12.7%), and VUS-Low (3,893; 84.1%) (**Fig. 4**).

Additionally, 74 CNV-positive samples classified as VUS underwent manual review for potential reclassification, of which 41 (55.4%) were reclassified as likely pathogenic. Overall 318 individuals were reclassified as carrying P/LP variants. This increased the overall diagnostic yield by approximately 1.4 percentage points. The diagnostic yield following reclassification across the different clinical categories is provided in **Table 2**.

**Table 2:** Re-classification of VUS samples diagnosis summary across clinical categories.

| **Cancer type** | **Total samples** | **Diagnosed samples(Samples with P/LP variants)** | **Diagnostic rate (%)** | **Newly added samples with P/LP variants** | **Diagnostic rate post-reclassification** | **Increase in diagnostic rate** |
|---|---|---|---|---|---|---|
| Breast | 10292 | 2092 | 20.3 | 124 | 21.5 | 1.2 |
| Ovarian | 3935 | 1245 | 31.6 | 42 | 32.7 | 1.1 |
| Colorectal | 1267 | 534 | 42.1 | 33 | 44.8 | 2.7 |
| Prostate | 762 | 66 | 8.7 | 12 | 10.2 | 1.5 |
| Endometrial | 539 | 120 | 22.3 | 9 | 23.9 | 1.6 |
| Asymptomatic | 2764 | 527 | 19.1 | 45 | 20.7 | 1.6 |
| Others | 3214 | 861 | 26.8 | 53 | 28.4 | 1.6 |

### Copy number variant detection by MLPA

MLPA was performed in 6,071 individuals, comprising 1,547 individuals who underwent MLPA testing alone and 4,524 who underwent combined NGS and MLPA testing. Among the MLPA-only group, 180 (11.63%) individuals were CNV positive, while 221 (4.88%) of those in the combined testing group were CNV positive. Within the combined testing cohort, NGS identified CNVs in 192 individuals (4.24%), while 251 individuals (5.54%) had CNVs detected by either NGS or MLPA. Of these, 162 individuals were positive by both methods, 59 were positive by MLPA but had no CNVs detected by NGS, and 30 were positive by NGS but negative by MLPA.

Among the 401 MLPA-positive individuals across both testing groups, the most frequently affected genes were *BRCA1* (118; 29.4%), followed by *PMS2* (60; 15.0%), *FANCA* (58; 14.5%), *MSH2* (32; 8.0%), *EPCAM* (22; 5.5%), *MLH1* (17; 4.2%), *BRCA2* (15; 3.7%), *PALB2* (14; 3.5%), *APC* (11; 2.7%), and *STK11* (10; 2.5%). Collectively, these 10 genes accounted for 357 (89.0%) of the MLPA-positive individuals, with *BRCA1* alone accounting for nearly one-third of the findings.

## Discussion

This retrospective analysis of 22,773 individuals represents one of the largest hereditary cancer datasets from India, highlighting substantial inherited cancer susceptibility, recurrent variants, and interpretation challenges across major cancer types.

The high diagnostic yield observed in colorectal cancer may partly reflect the substantial representation of early-onset disease in our cohort. The mean age at testing was 50.3 ± 15.4 years, and 46.5% of patients were diagnosed at or before 50 years of age. P/LP variants in high-penetrance susceptibility genes, including *MLH1*, *MSH2*, and *APC* were more frequently observed in this age group, supporting a substantial burden of hereditary predisposition among younger patients. The clinically selected nature of the cohort may also have contributed to the observed yield. In breast cancer, the diagnostic yield was comparable to that reported in Indian cohorts meeting National Comprehensive Cancer Network (NCCN) testing criteria (approximately 18%–30%)^18–21,37,38^. This consistency suggests that our cohort reflects a similar burden of hereditary predisposition despite potential differences in selection strategies.

Age-stratified analysis showed that diagnostic yield generally decreased with increasing age in breast, colorectal, ovarian, and endometrial cancers, whereas prostate cancer showed an increasing trend with age. The highest yields for colorectal, ovarian, and endometrial cancers were observed in the 30–49-year age group. These findings highlight the importance of considering both age and family history while recognizing that their associations with germline variant detection may differ across cancer types. Consistent with this, Stadler *et al*. reported that restricting germline testing to patients diagnosed before age 50 would miss a substantial proportion of pathogenic variant carriers^39^. However, the age-related trends observed in endometrial and prostate cancers should be interpreted in light of the smaller subgroup sizes and wider confidence intervals. Family history-based analysis showed that a family history of the corresponding cancer was associated with higher diagnostic yields across most major cancer types. However, the majority of individuals with P/LP variants had no reported family history of the corresponding cancer, indicating that family-history-based testing alone may miss a substantial proportion of hereditary cancer cases. Multi-variate analysis further demonstrated that both age and family history are independently associated with diagnostic yield.

We also observed differences in diagnostic yield according to metastatic status. The diagnostic yields were generally higher in non-metastatic cases, with ovarian cancer being the exception. These findings highlight the importance of considering cancer-specific clinical context when interpreting germline testing results. The clinical relevance of metastatic presentation is further underscored by a recent study of 76,356 women from India’s National Cancer Registry Programme, which reported that 12.96% of patients with breast cancer presented with metastatic disease at diagnosis, highlighting the burden of advanced-stage disease in the Indian population^40^.

Consistent with earlier Indian multigene panel studies, *BRCA1* and *BRCA2* accounted for a substantial proportion of P/LP variants. *BRCA1* pathogenic variants were predominant over *BRCA2* in Indian patients, corroborating previous reports from Indian cohorts^18–20,22^. Differences in *BRCA1/2* variant composition and spectrum have been reported across Indian, Chinese, Korean, and Japanese populations, with more than half of the identified variants being Asian-specific, highlighting the ethnic specificity of *BRCA1/2* variation within Asia^41^.

A notable finding in our cohort was the recurrence of the BRCA1 frameshift variant c.68_69del (p.Glu23ValfsTer17), identified in 351 individuals. Although originally described as an Ashkenazi Jewish founder mutation, this variant has also been reported in Indian populations, with evidence suggesting regional enrichment in South India^18–21,37,38^. Its recurrence in our cohort raises the possibility of a population-specific founder effect, although further haplotype-based and population-level studies are required to establish this. Other recurrent *BRCA1* variants, including p.Arg1203Ter and p.Ser1503Ter, further emphasize the importance of population-specific variant characterization. In addition, the strong concordance between the P/LP findings and functional evidence from BRCA1 saturation genome-editing studies supports the utility of functional datasets in variant interpretation. In this context, the BRCAIndica compendium provides a valuable population-specific resource for interpreting BRCA1/2 variants reported in India^42^.

Beyond *BRCA1/2*, P/LP variants were identified in other homologous recombination repair genes, including *ATM*, *PALB2*, *CHEK2*, *RAD51C*, *RAD51D*, and *BRIP1*, demonstrating the value of multigene panel testing in patients with breast and/or ovarian cancer^18^. International studies have also demonstrated associations between pathogenic variants in genes such as *BRCA1*, *BRCA2*, *PALB2*, *CHEK2*, and *ATM* and breast cancer risk, with penetrance influenced by family history and other epidemiological factors^43–47^. Large multigene testing studies involving women with breast cancer have further reported racial and ethnic differences in the prevalence of pathogenic variants across susceptibility genes and in gene-specific breast cancer risk associations, highlighting the importance of considering population background in hereditary breast cancer risk assessment^48^.

The importance of characterizing rare, cancer-specific susceptibility variants is illustrated by a large study of more than 3.3 million individuals, which reported a strong association between germline EGFR T790M and lung cancer risk (OR: 25.18), with a possible founder effect in Southern Appalachian populations^49^. Although this variant was not identified in our cohort, such findings emphasize the importance of investigating rare susceptibility variants across diverse populations. We also identified the recently described Indian founder variant MLH1 c.306G>T in 25 individuals, supporting its contribution to Lynch syndrome in the Indian population^50,51^. Its detection, together with the substantial representation of early-onset colorectal cancer in our cohort, highlights the importance of systematic germline evaluation for hereditary colorectal and endometrial cancer susceptibility.

Breast cancer subtype analysis showed that ER−/HER2− was the most common subtype, accounting for 59.2% of cases, and had the highest diagnostic yield for P/LP variants (25.5%; O/E ratio: 1.3). *BRCA1* accounted for 66.8% of P/LP variants in this subgroup. Previous studies have reported a substantial burden of triple-negative breast cancer in Indian patients^52,53^. The high proportion of P/LP variants observed in the ER−/HER2− subgroup in our cohort further highlights the relevance of tumor subtype in assessing hereditary cancer susceptibility.

Reanalysis of VUS using the in-house automated ACMG tool, followed by manual review, confirmed 277 SNV/indel-positive samples as P/LP. An additional 41 of 74 CNV-positive samples classified as VUS were reclassified as likely pathogenic following manual review. Collectively, these reclassifications increased the overall diagnostic yield by 1.4 percentage points. The remaining VUS were further stratified into VUS-High, VUS-Mid, and VUS-Low categories using the *VUS Grader* tool. These findings demonstrate the potential clinical value of periodic variant reinterpretation as new classification evidence becomes available. However, variant interpretation in populations underrepresented in genomic reference datasets remains challenging. An ancestry-specific analysis of 9,899 TCGA cancer cases found that South Asian individuals accounted for only 0.5% of the cohort, with most predisposing variants observed exclusively in a single ancestry, highlighting the need for larger, more diverse genomic datasets and population-informed variant interpretation^54^.

NGS and MLPA provided complementary information for CNV detection. In the combined testing cohort of 4,524 individuals, 251 had CNVs detected by either method. Notably, 59 individuals had CNVs detected by MLPA but not by NGS, whereas 30 had CNVs detected by NGS but not by MLPA. These findings support the use of complementary methods to improve CNV detection. In addition to the primary cohort, 1,547 samples underwent MLPA testing alone, of which 180 (11.63%) were positive, compared with 221 (4.88%) in the combined testing cohort. The higher positivity rate in the MLPA-only group may reflect referral enrichment, potentially due to prior clinical suspicion or the need for confirmatory CNV assessment. Therefore, these additional samples should be interpreted separately and not considered representative of CNV prevalence in the broader study population.

Taken together, our findings highlight the substantial and heterogeneous burden of germline cancer susceptibility across major cancer types in this Indian testing cohort. The cancer-specific associations with age and family history, recurrent and population-relevant variants, and additional diagnostic findings from VUS reinterpretation and complementary CNV testing support comprehensive germline assessment.

### Limitations of the study

This study has several limitations. First, the cohort comprised individuals referred for germline genetic testing by treating clinicians, introducing the possibility of referral bias. Consequently, the study population may be enriched for individuals with clinical features suggestive of hereditary cancer predisposition, and the diagnostic yields reported here may not be generalizable to unselected cancer populations. Second, clinical and phenotypic information was obtained primarily from test requisition forms and information provided by referring clinicians, without independent verification. In particular, family history information may have been based on patients’ verbal reports to their clinicians and was not independently verified. Similarly, hormone receptor status was obtained from test requisition forms and was not independently validated. Given the retrospective design and the use of clinical information available at the time of testing, independent validation of these variables was beyond the scope of this study.

## Conclusions

This large South Asian study provides insights into the germline variant landscape across major cancer types, highlighting the burden and genetic heterogeneity of hereditary cancer susceptibility in the Indian population. The variation in diagnostic yield across cancer types, identification of recurrent pathogenic variants, and complementary contributions of NGS and MLPA to CNV detection underscore the value of comprehensive germline testing. These findings support broader testing strategies, population-specific variant characterization, and periodic variant reinterpretation to improve the identification and assessment of inherited cancer susceptibility. Further characterization of hereditary cancer susceptibility across India’s diverse populations, together with improved representation in genomic reference datasets, will be important for advancing population-informed variant interpretation and hereditary cancer risk assessment in South Asia.

## Ethics statement

This retrospective, non-interventional study was conducted using existing de-identified patient data. Ethical approval was obtained from the MedGenome Lab’s Institutional Ethics Committee (IEC No. EC/NEW/INST/2022/3194, proposal No. 01/2023). All study procedures were conducted in accordance with the ethical principles of the Declaration of Helsinki and the ICMR’s National Ethical Guidelines for Biomedical and Health Research Involving Human Participants.

## Data Availability

All data produced in the present work are contained in the manuscript

## Acknowledgments

The authors sincerely thank Dr. Rajkumar Thangarajan for providing valuable inputs and critical feedback on the manuscript.

## Supplementary Figures

**Supplementary Figure 1.**
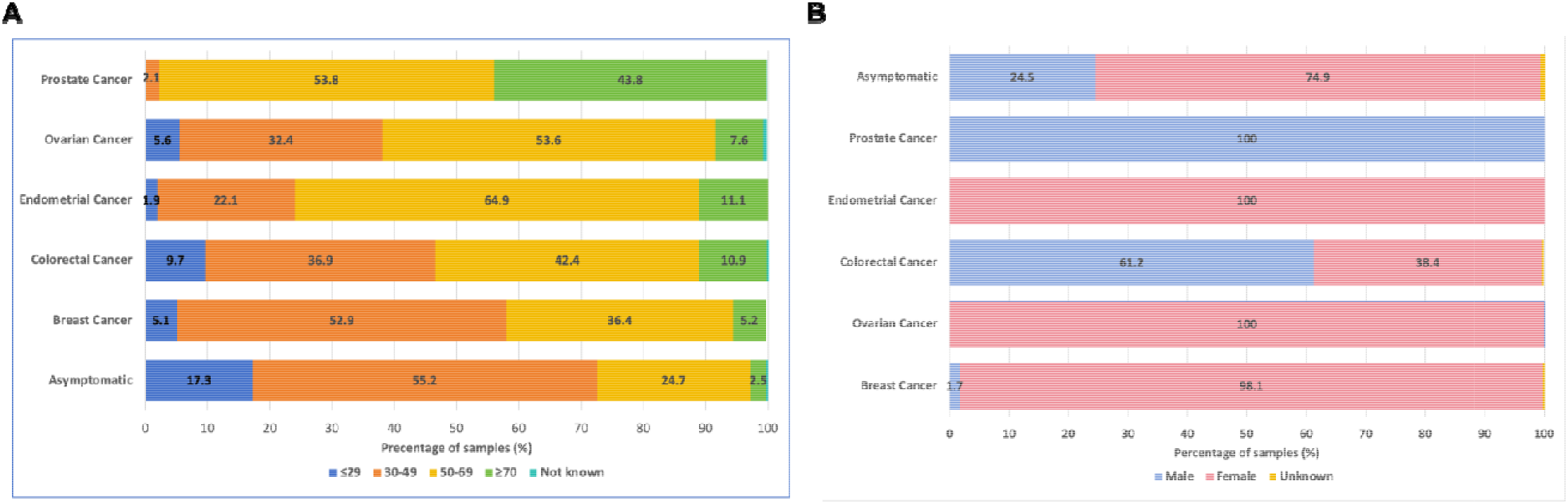
Age and sex distribution across cancer types. **(A)** Age distribution across cancer types and the asymptomatic group. The x-axis represents age groups, and the y-axis represents cancer types and the asymptomatic group. **(B)** Sex distribution across cancer types and the asymptomatic group. The x-axis represents the percentage of individuals, and the y-axis represents cancer types and the asymptomatic group.

**Supplementary Figure 2.**
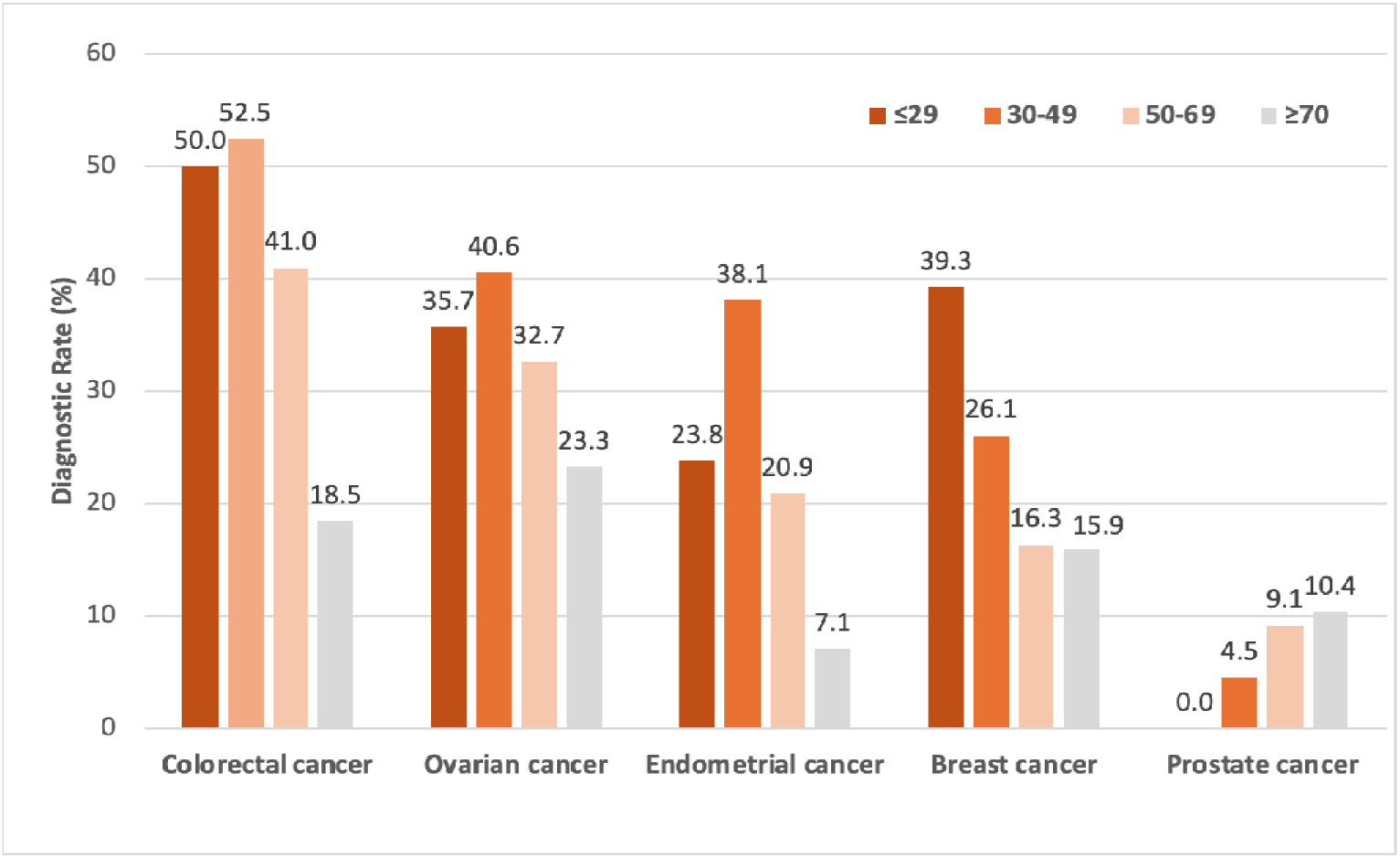
Age group-wise diagnostic yield across the five major cancer types. The diagnostic yield (%) across colorectal, ovarian, endometrial, breast, and prostate cancers is shown for four age groups (≤29, 30–49, 50–69, and ≥70 years). The x-axis represents the major cancer types, and the y-axis represents the diagnostic yield (%). Age-group classifications are indicated in the legend.

**Supplementary Figure 3.**
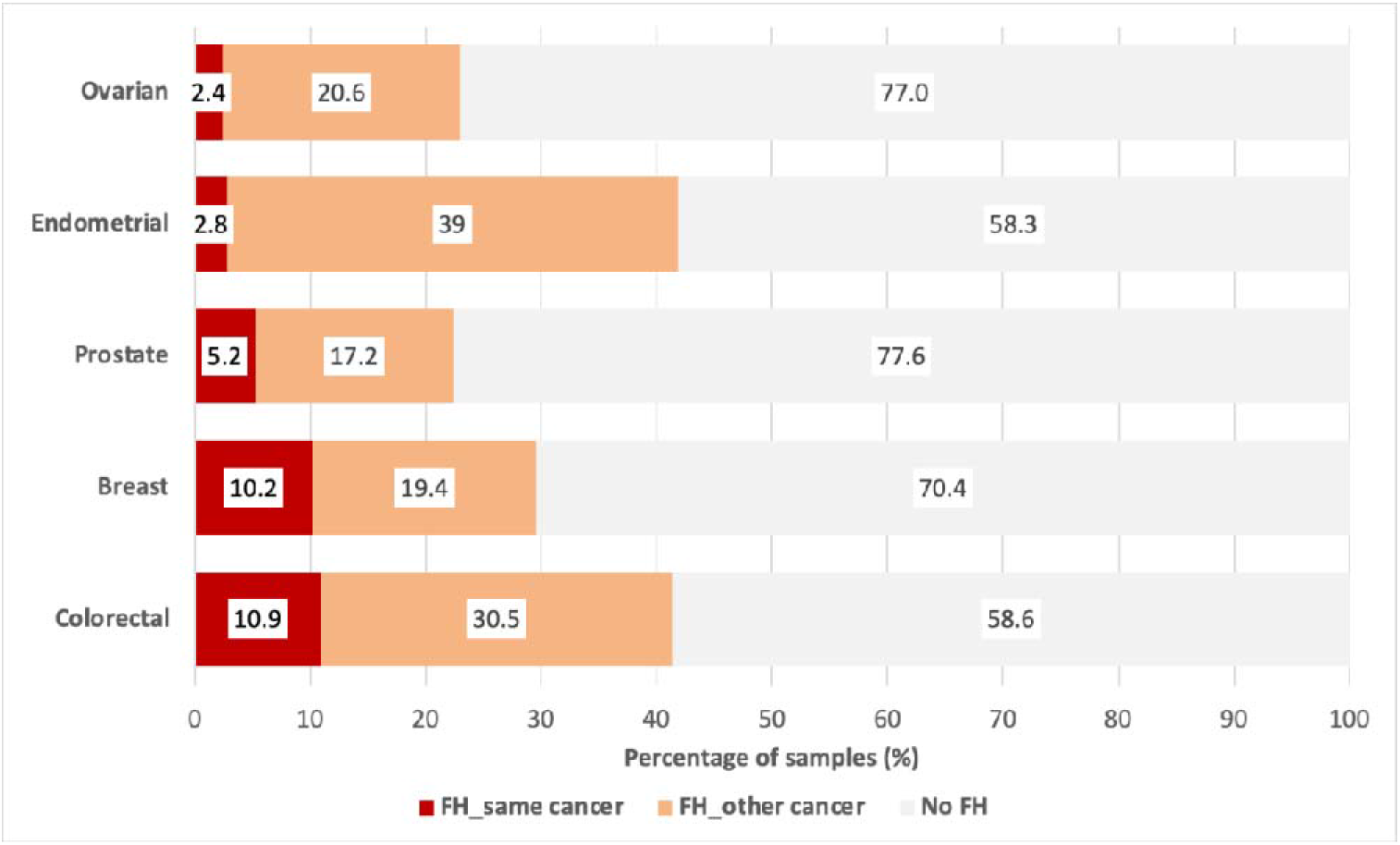
Family history patterns across the five major cancer types. The horizontal stacked bar chart shows the percentage of individuals reporting a family history of the corresponding cancer (FH_same cancer), a family history of other cancer types (FH_other cancer), or no family history of cancer (No FH) across colorectal, breast, prostate, endometrial, and ovarian cancer groups.

**Supplementary Figure 4.**
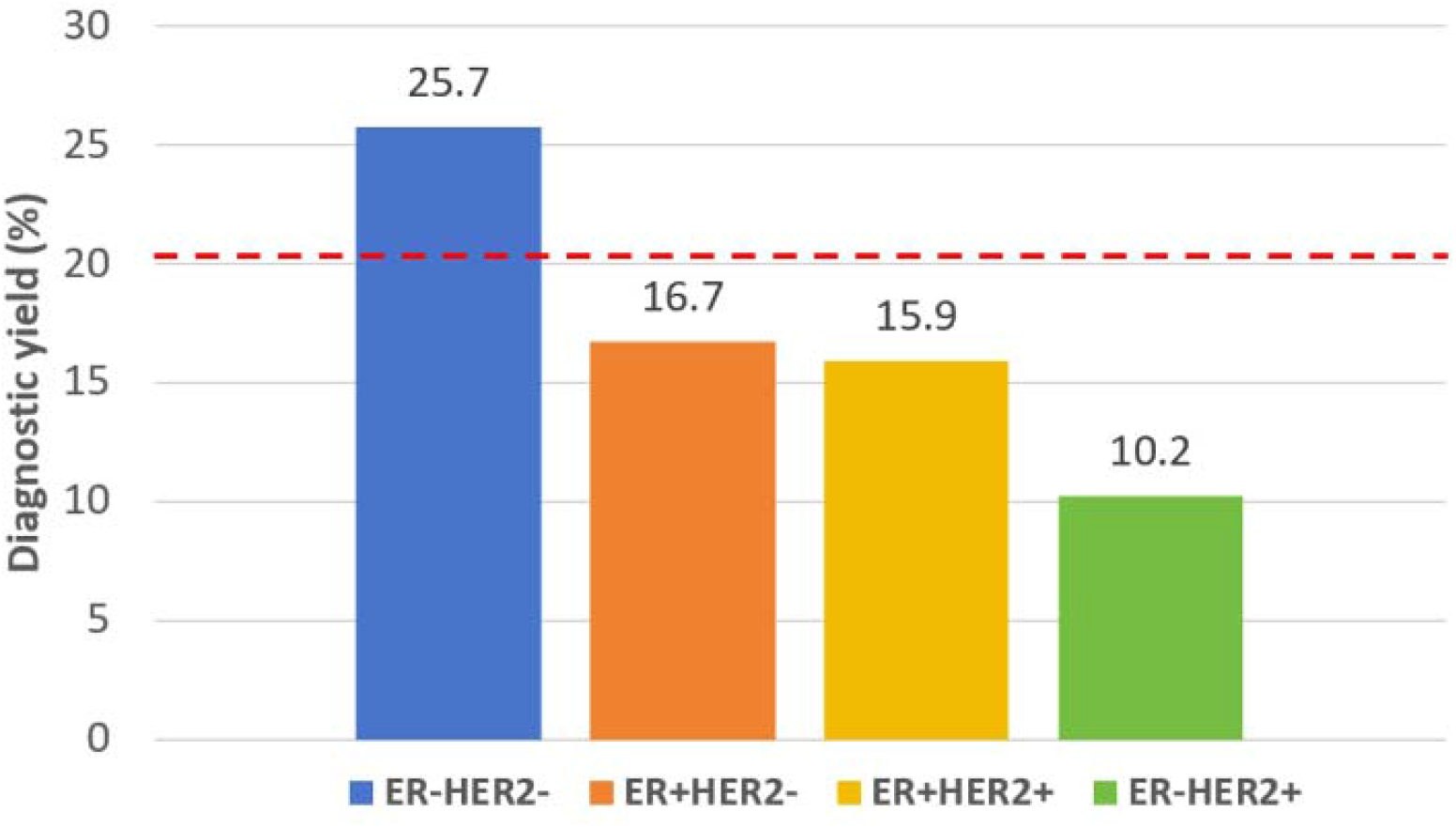
Diagnostic yield across hormone receptor-based breast cancer subtypes. Bars represent the diagnostic yield (%) of pathogenic or likely pathogenic (P/LP) variants across the four subgroups defined by estrogen receptor (ER) and human epidermal growth factor receptor 2 (HER2) status. The x-axis represents breast cancer subtypes, and the y-axis represents diagnostic yield (%). The red dashed line indicates the overall diagnostic yield in the breast cancer cohort (20.3%).

